# Estimating the population-level kidney benefits of improved uptake of SGLT2 inhibitors in patients with chronic kidney disease in Australian primary care

**DOI:** 10.1101/2023.06.26.23291881

**Authors:** Brendon L Neuen, Min Jun, James Wick, Sradha Kotwal, Sunil V Badve, Meg J Jardine, Martin Gallagher, John Chalmers, Kellie Nallaiah, Vlado Perkovic, David Peiris, Anthony Rodgers, Mark Woodward, Paul E Ronksley

## Abstract

**Background:** Although sodium glucose co-transporter 2 (SGLT2) inhibitors reduce the risk of kidney failure and death in patients with chronic kidney disease (CKD), they are underused in routine clinical practice. We evaluated the number of patients with CKD in Australia that would be eligible for treatment with an SGLT2 inhibitor and estimated the number of cardiorenal and kidney failure events that could be averted with improved uptake of SGLT2 inhibitors.

**Methods:** Using nationally-representative Australian primary care data (MedicineInsight), we identified patients that would have met inclusion criteria of the CREDENCE, DAPA-CKD, and EMPA-KIDNEY trials between 1 January 2020 and 31 December 2021. We applied these data to age and sex-stratified estimates of CKD prevalence from the broader Australian population (using national census data) to generate population-level estimates for: (1) the number of CKD patients eligible for treatment with SGLT2 inhibitors and (2) the annual number of potentially preventable cardiorenal (CKD progression, kidney failure, or death due to cardiovascular disease or kidney failure), and kidney failure events with SGLT2 inhibitors based on trial event rates.

**Results:** In MedicineInsight, 44.2% of adults with CKD would have met CKD eligibility criteria for an SGLT2 inhibitor; baseline use was 4.1%. Applying these data to the broader Australian population, we estimated 230,246 patients with CKD in Australia would have been eligible for treatment with any SGLT2 inhibitor. Optimal implementation of SGLT2 inhibitors (75% uptake in eligible patients) could reduce cardiorenal and kidney failure events annually in Australia by 3,644 (95% CI 3,526-3,764) and 1,312 (95% CI 1,242-1,385), respectively.

**Conclusions:** Improved uptake of SGLT2 inhibitors for patients with CKD in Australian primary care has the potential to prevent large numbers of patients experiencing CKD progression or dying due to cardiovascular or kidney disease. Identifying strategies to increase the uptake of SGLT2 inhibitors is critical to realising the population-level benefits of this drug class.

## Introduction

Sodium glucose cotransporter 2 (SGLT2) inhibitors reduce the risk of chronic kidney disease (CKD) progression by approximately 40%, regardless of diabetes, primary kidney diagnosis or kidney function and are, therefore, now considered foundational therapy for CKD alongside renin-angiotensin system (RAS) blockade in major clinical practice guidelines.^1–4^

Despite clear evidence from multiple large, randomised trials that SGLT2 inhibitors reduce the risk of kidney failure or death in people with CKD, the uptake of these agents in routine practice remains low. In the United Kingdom and United States, only about 10% of patients with diabetes and CKD are prescribed an SGLT2 inhibitor; paradoxically, those at higher risk of kidney failure are less likely to receive one.^5^ An almost identical phenomenon is observed for adjacent disease states, such as type 2 diabetes with atherosclerotic cardiovascular disease and heart failure (regardless of diabetes), whereby uptake of SGLT2 inhibitors has been limited and those at highest risk are less likely to receive one.^6^ The explanation for the underuse of SGLT2 inhibitors in routine practice is complex and not fully understood but highlights a major opportunity to improve population health if the uptake of SGLT2 inhibitors is improved.

Understanding the potential population-level benefits of SGLT2 inhibitors in patients with CKD might encourage policymakers, health systems and clinicians to improve access and increase the use of SGLT2 inhibitors. Therefore, using routinely collected primary care data in Australia, we sought to evaluate the number of patients with CKD who would be eligible for treatment with an SGLT2 inhibitor, based on inclusion and exclusion criteria of SGLT2 inhibitor kidney outcome trials, and estimate the annual number of cardiorenal events and patients progressing to kidney failure that could be averted at the population-level with improved uptake of SGLT2 inhibitors in patients with CKD.

## Methods

This study abides by the Strengthening the Reporting of Observational Studies in Epidemiology (STROBE) reporting guidelines for observational studies.^7^

### Data sources

We used routinely collected data from MedicineInsight, a nationally-representative primary care data source. MedicineInsight includes de-identified patient information, clinical data and comorbidities, prescription medications and laboratory test results (including estimated glomerular filtration rate [eGFR] and urine albumin:creatinine ratio [UACR]) on patients visiting primary care practices participating in MedicineInsight from all states and territories in Australia. Detailed information about MedicineInsight has been previously published.^8, 9^ This study was approved by the MedicineInsight Data Governance Committee (2020-004) and Research Ethics Review Committee of the Sydney Local Health District, NSW, Australia (X21-0428, 2020/ETH00963).

### Population

Among a cohort of all adults (age ≥18 years) with ≥1 clinical encounter and ≥1 eGFR measurement (with or without a UACR measurement) between 1 January 2020 and 31 Dec 2021 (n=1,217,398), we identified patients with eGFR <60 mL/min/1.73m^2^ and/or UACR >30 mg/g. These patients were categorised into three groups based on the eGFR and UACR eligibility criteria of three SGLT2 inhibitor kidney outcome trials: CREDENCE, DAPA-CKD and EMPA-KIDNEY.^11–13^

For each SGLT2 inhibitor trial-based group, the index date was defined as the date of the qualifying eGFR or UACR value in MedicineInsight during the study period (i.e., the first date during the two-year study period on which the eGFR or UACR inclusion criteria of the SGLT2 inhibitor trial was met). We applied key exclusion criteria from each trial that could be defined using data from MedicineInsight (Table S1), except for maximum tolerated or labelled dose of RAS blockade to better reflect real world practice. Identification of the study cohorts is displayed in Figure S1.

We used age- and sex-stratified estimates of CKD prevalence derived from MedicineInsight^14^ mapped to corresponding population census data in 2021 from the Australian Bureau of Statistics^15^ to estimate the number of prevalent patients with CKD in Australia based on conservative, moderate and high-estimate models (Table S2). The conservative model (CKD defined as two consecutive eGFR measurements <60 mL/min/1.73m^2^, ≥90 days apart, and/or two consecutive UACR >30mg/g, ≥90 days apart) was used in the main analysis with moderate and high estimate models (defined in Table S2) used as sensitivity analyses.

### Outcomes

The primary outcome in this study was a composite cardiorenal outcome that included CKD progression (sustained 40%, 50% or 57% decline in eGFR), kidney failure or death due to cardiovascular disease or kidney failure, based on the primary outcomes of the three SGLT2 inhibitor outcome trials.

Secondary outcomes included a kidney composite outcome that was defined as the primary outcome excluding cardiovascular death, and kidney failure alone. Kidney failure was defined as maintenance dialysis, kidney transplantation or a sustained low eGFR (<15 mL/min/1.73m^2^ in CREDENCE and DAPA-CKD, and <10 mL/min/1.73m^2^ in EMPA-KIDNEY). Because CKD progression was defined using different eGFR decline thresholds across each of the trials, we also conducted a sensitivity analysis using a standardised kidney composite outcome of 40% decline in eGFR, kidney failure or death due to kidney failure.^16^ Ketoacidosis was also evaluated as the main recognised serious adverse effect of SGLT2 inhibitor use.^1^

### Statistical Analysis

We computed incidence rates, expressed as the number of events per 1000 patient-years, in SGLT2 inhibitor and placebo groups for key outcomes in each trial, based on published reports. Applying relative treatment effects from each trial to the placebo event rate for key outcomes, we estimated absolute risk reductions (ARR) and numbers-needed-to-treat (NNT) over 1 and 3 years, as has been done in previous work estimating the population-level benefits of SGLT2 inhibitors.^17, 18^ Next, we estimated the annual number of potentially preventable events in Australia (or potential ketoacidosis events caused) with implementation of SGLT2 inhibitors by applying these event rates to national estimates of the total number of CKD patients eligible for SGLT2 inhibitors. We defined optimal implementation as 75% uptake of SGLT2 inhibitors among eligible patients and explored the effect of varying levels of treatment uptake (25%, 50%, 75% and 100%) on the number of potentially preventable events. Finally, we estimated the number of potentially preventable events with optimal implementation of SGLT2 inhibitors across major subsets of patients with CKD by applying trial event rates from key subgroups – diabetes (yes/no), eGFR (<45 and ≥45 mL/min/1.73m^2^) and UACR (≤300 and >300 mg/g).

All analyses were done using Stata version 17 (StataCorp LLC, College Station, TX).

## Results

In MedicineInsight, we identified 147,110 (12.1%) adult patients who had an eGFR <60 mL/min/1.73m^2^ and/or UACR >30 mg/g between 1 January 2020 and 31 December 2021. After applying key trial-specific inclusion and exclusion criteria (Table S1 and S3), 11,046 (7.5%) of 147,110 patients in MedicineInsight were eligible for CREDENCE, 25,455 (17.3%) were eligible for DAPA-CKD, and 65,049 (44.2%) eligible for EMPA-KIDNEY.

Baseline characteristics of patients with CKD eligible for treatment with an SGLT2 inhibitor in MedicineInsight and randomised trials are displayed in Table 1. Compared to randomised trials, eligible patients in MedicineInsight were typically older, more likely to be female, have higher eGFR, and less likely to have diabetes (apart from CREDENCE which exclusively enrolled patients with diabetes) and cardiovascular disease (except EMPA-KIDNEY). Median UACR was similar for EMPA-KIDNEY eligible patients, but lower for patients eligible for CREDENCE and DAPA-CKD. The use of RAS blockade was considerably lower in MedicineInsight (approximately 65%) compared to the SGLT2 inhibitor trials in which almost all patients were receiving RAS blockade, as mandated for entry into the trials. Use of SGLT2 inhibitors in MedicineInsight ranged from 4.1% in EMPA-KIDNEY eligible patients to 14.4% in CREDENCE eligible patients.

**Table 1.**
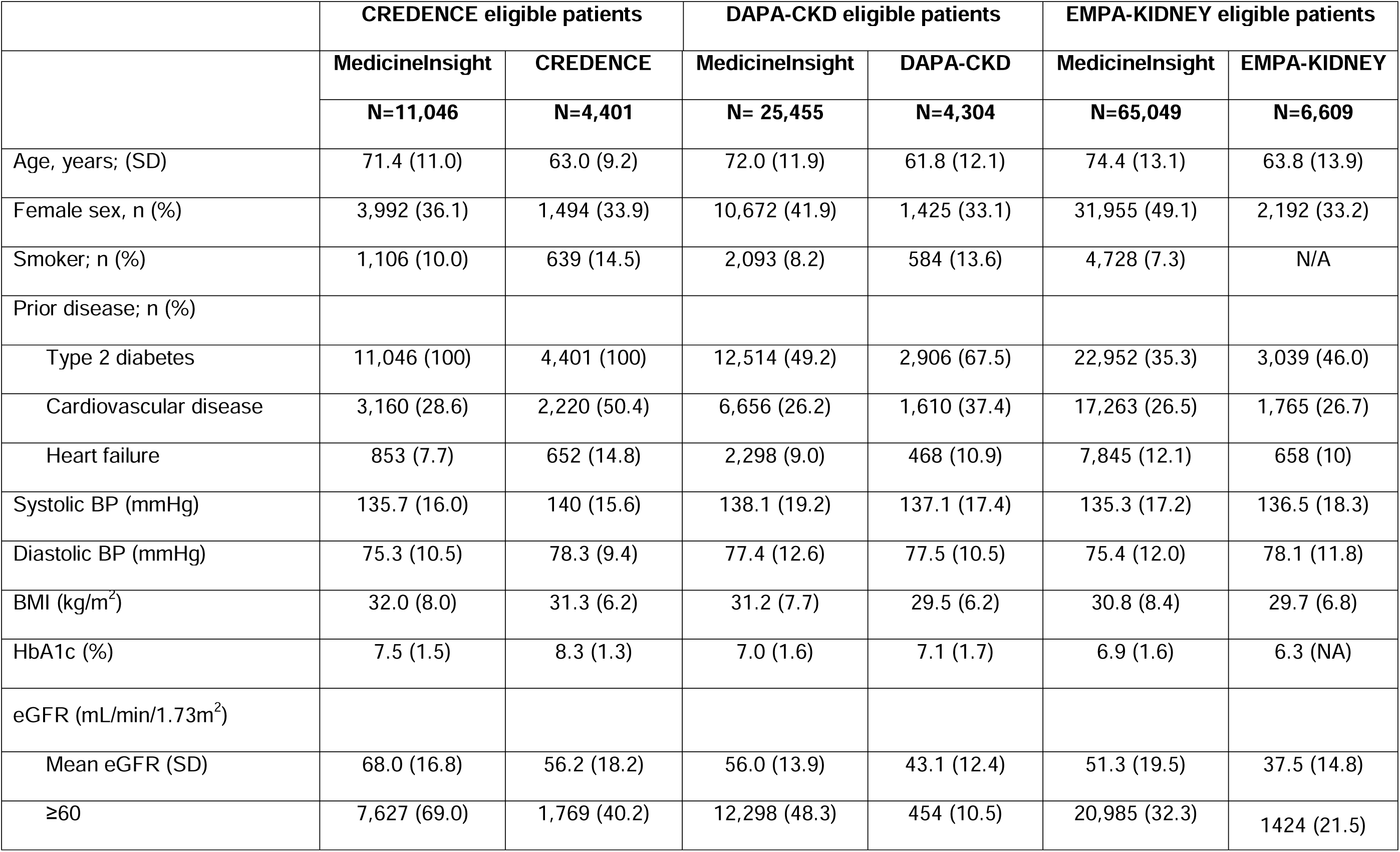
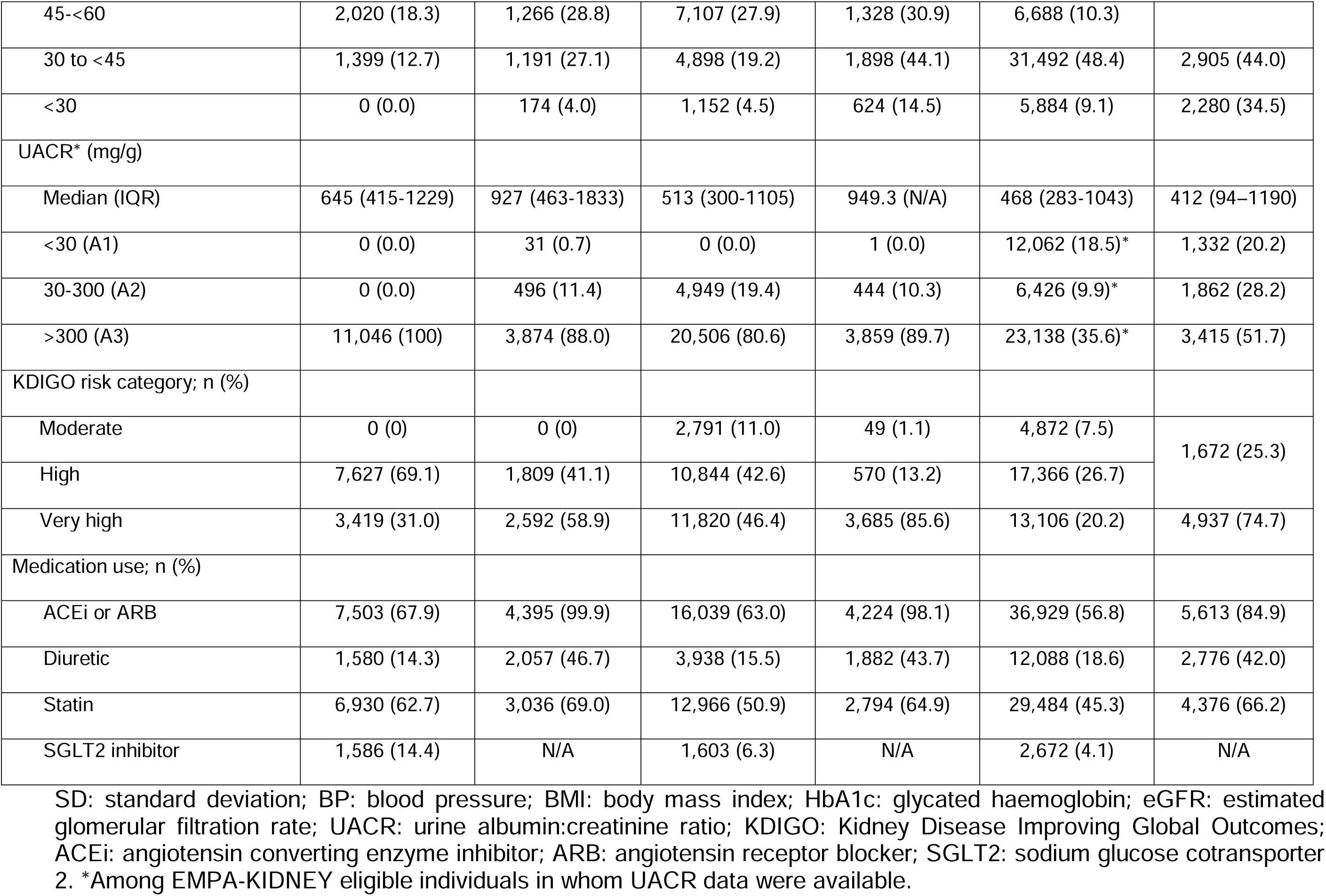
Baseline characteristics of patients with CKD eligible for treatment with an SGLT2 inhibitor in MedicineInsight (2020-2021) and SGLT2 inhibitor kidney outcome trials

Applying the proportion of CKD patients in MedicineInsight eligible for SGLT2 inhibitor treatment to the broader Australia population based on a conservative model of CKD prevalence in Australia, an estimated 39,098, 90,100 and 230,246 patients would have been eligible for CREDENCE, DAPA-CKD and EMPA-KIDNEY, respectively (Table S4). EMPA-KIDNEY had the broadest inclusion criteria of all three SGLT2 inhibitor trials, encompassing the eGFR and UACR criteria for the other trials, therefore representing patients eligible for treatment with any SGLT2 inhibitor (Figure 1; Table S5).

**Figure 1.**
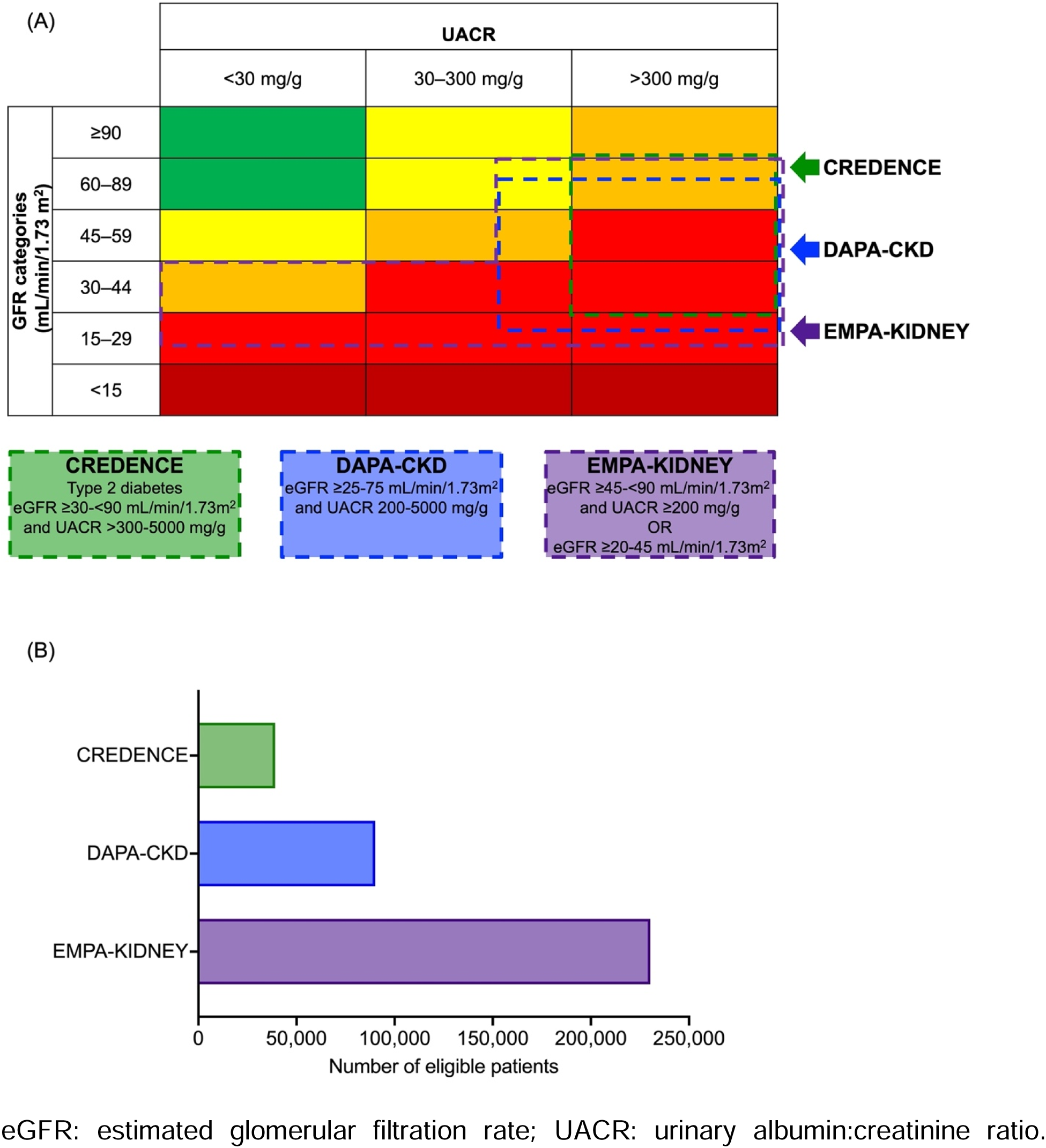
(A) eGFR and UACR inclusion criteria in SGLT2 inhibitor kidney outcome trials and (B) number of Australian primary care patients estimated to be eligible for treatment with an SGLT2 inhibitor.

Event rates, relative treatment effects, ARR and NNTs over 1 and 3 years for key outcomes in each trial are displayed in Figure 2. For the primary cardiorenal outcome, absolute risk reductions at 3 years ranged from 4.7-6.0% with corresponding NNTs of 14-21. Absolute risk reductions for kidney failure alone at 3 years were 2.2-3.7% with NNTs of 15-27. In sensitivity analyses, ARR and NNTs at 1 and 3 years for the standardised kidney outcome were similar to the main kidney composite outcome (Table S6).

**Figure 2.**
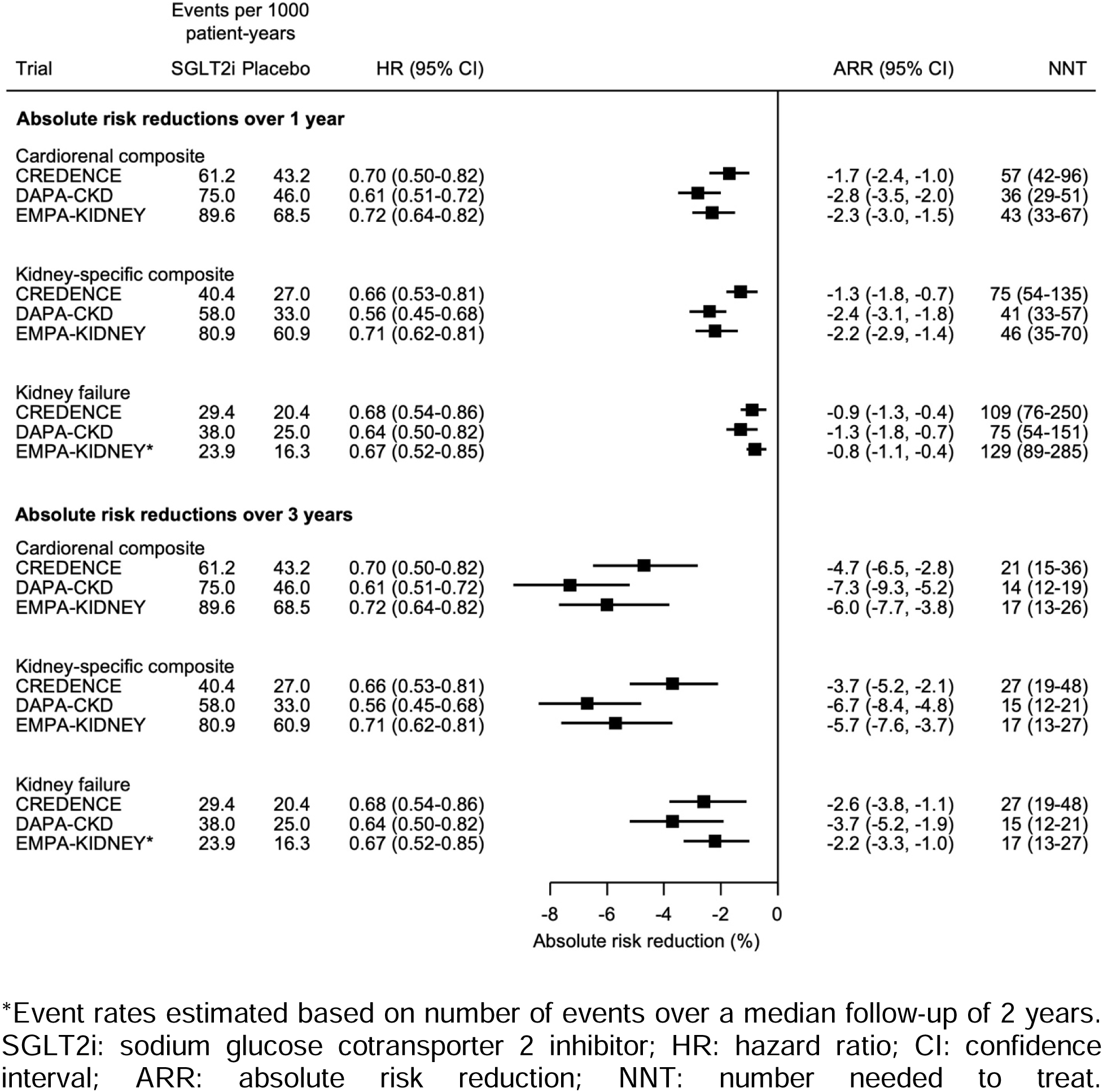
Treatment effects, absolute risk reductions and numbers-needed-to-treat at 1 and 3 years in SGLT2 inhibitor kidney outcome trials.

Estimates of the annual number of events potentially prevented (or ketoacidosis events caused) in Australia with optimal implementation of SGLT2 inhibitors in CKD are displayed in Figure 3. Optimal implementation of SGLT2 inhibitors in eligible patients with CKD is projected to result in 3,644 fewer cardiorenal events (95% CI 3,526-3,764), 3,454 fewer kidney-composite events (95% CI 3,339-3,571) and 1,312 fewer (95% CI 1,242-1,385) patients progressing to kidney failure annually. Estimated annual reductions for the standardised kidney-composite outcome are displayed in Figure S2. The absolute benefits for these outcomes substantially outweighed the risk of ketoacidosis. Estimated number of events potentially prevented or caused in MedicineInsight, and nationally using three models of CKD prevalence are displayed in Table S7. Figure 4 displays the number of potentially preventable events in Australia with varying levels of SGLT2 inhibitor uptake; even modest increases in SGLT2 inhibitor use (i.e., 25% uptake) may result in 437 fewer patients (95% CI 397-480) in Australia reaching kidney failure annually.

**Figure 3.**
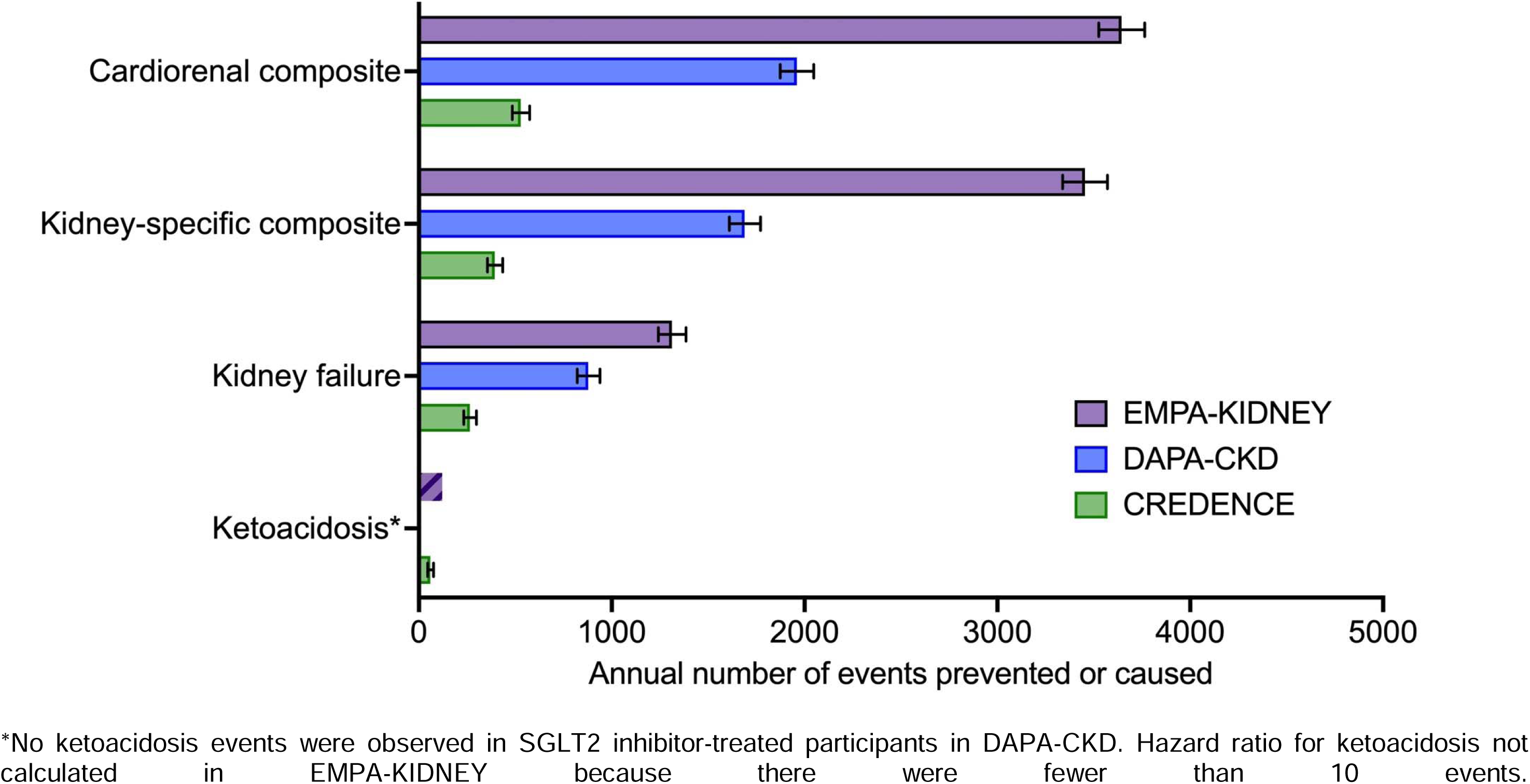
Estimated annual number of events expected to be prevented or caused in Australia with optimal implementation (75% uptake) of SGLT2 inhibitors in patients with CKD

**Figure 4.**
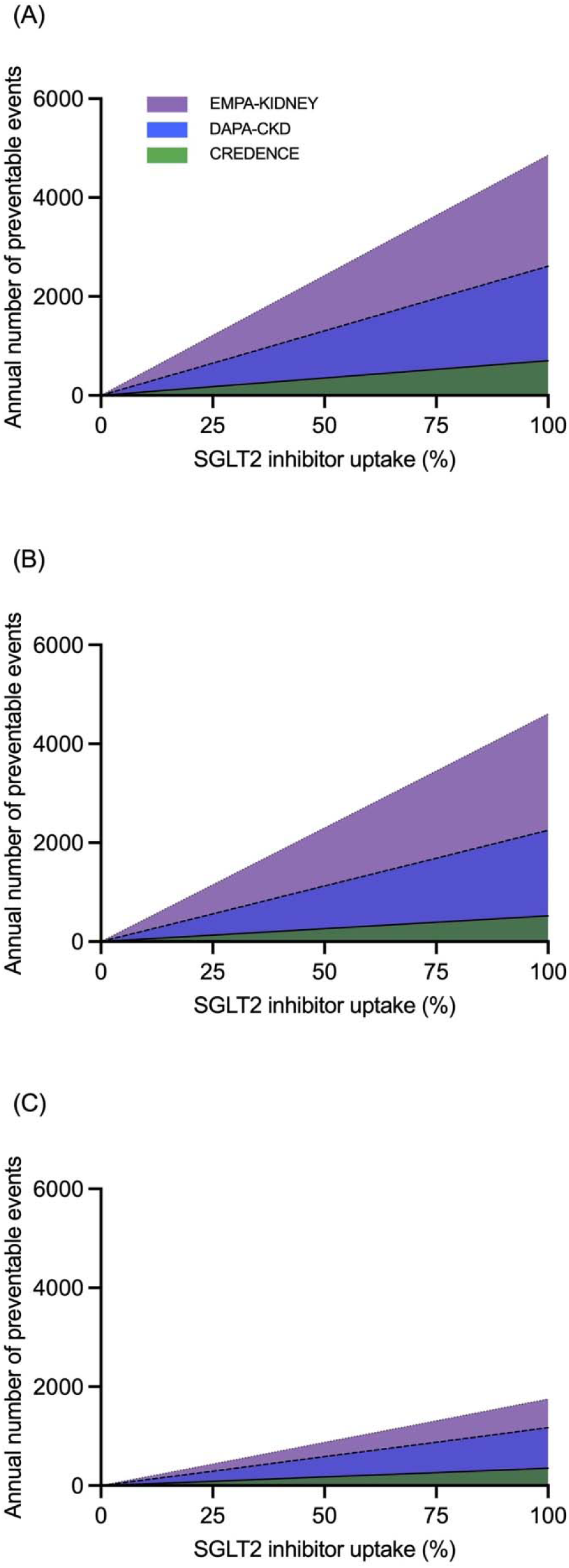
Estimated annual number of (A) cardiorenal, (B) kidney-specific composite and (C) kidney failure events expected to be prevented in Australia with different levels of SGLT2 inhibitor uptake in patients with CKD.

Implementation of SGLT2 inhibitors is projected to result in fewer cardiorenal events in patients with and without diabetes and those with eGFR <45 and ≥45 mL/min/1.73m^2^ (Figure 5). Because patients with UACR ≤300 mg/g were either excluded (CREDENCE) or represented in relatively small numbers in SGLT2 inhibitor kidney outcome trials, cardiorenal events prevented were predominantly observed for patients with UACR >300 mg/g compared to those with UACR ≤300 mg/g. Number of events, event rates, and treatment effects across subgroups in each trial are displayed in Table S8. Estimated cardiorenal events potentially prevented in MedicineInsight, and nationally using three models of CKD prevalence are displayed in Table S9.

**Figure 5.**
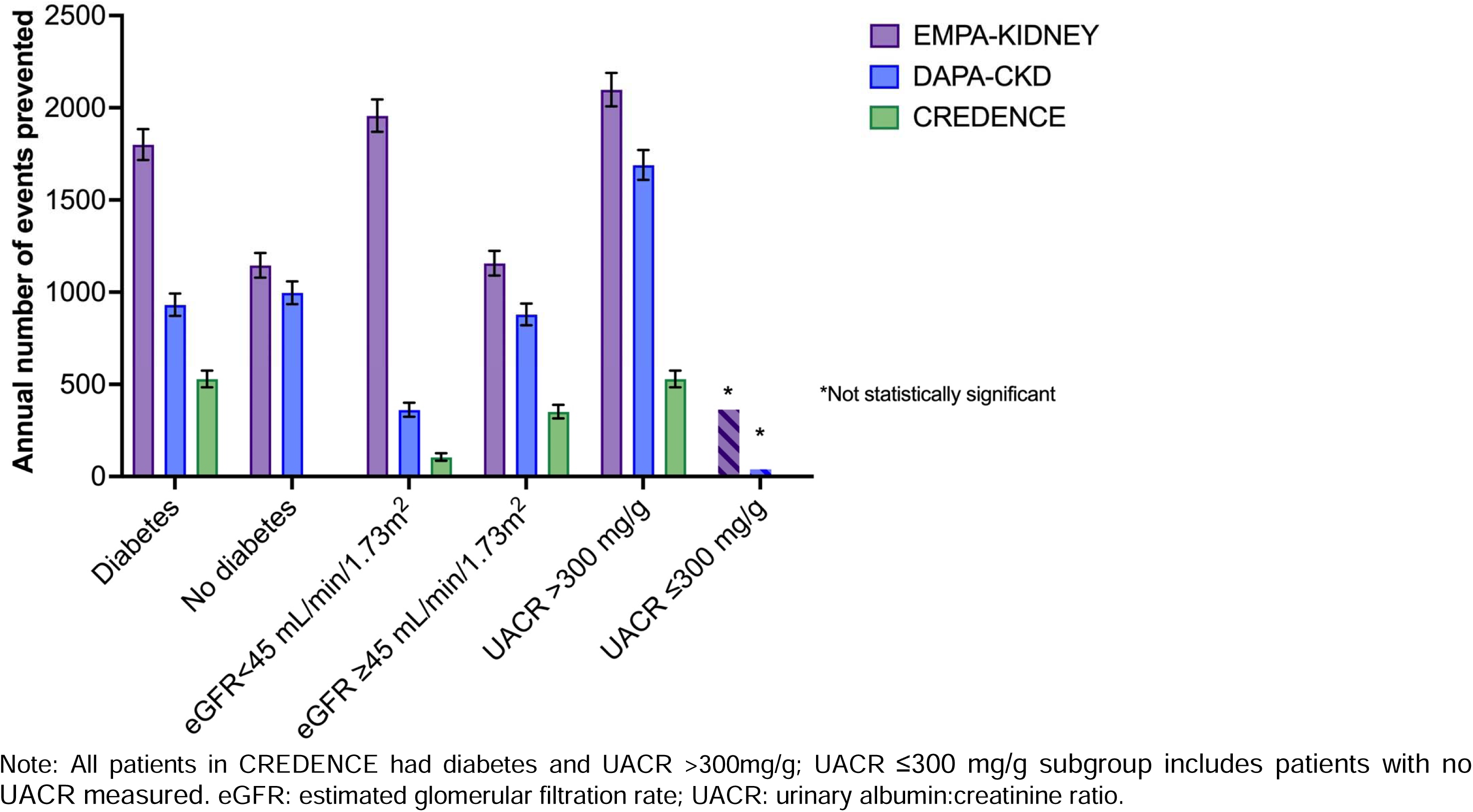
Estimated annual number of cardiorenal events expected to be prevented in Australia with optimal implementation (75% uptake) of SGLT2 inhibitors in patients with CKD according to diabetes status, eGFR and UACR.

## Discussion

In this observational study using a nationally-representative primary care data source from Australia, we observed that a substantial number of patients with CKD in Australian primary care may be eligible for treatment with an SGLT2 inhibitor, based on trial inclusion criteria. The number and proportion of patients eligible for treatment varied based on inclusion criteria of each trial, with nearly 45% of patients with CKD eligible for treatment with any SGLT2 inhibitor. Our estimates suggest that increased uptake of SGLT2 inhibitors in CKD could translate to significant population-level benefits including preventing large numbers of people experiencing CKD progression, kidney failure or death due to cardiovascular disease or kidney failure annually, as well as substantial reductions in the number of patients reaching kidney failure. These benefits outweighed the relatively small absolute increased risk of ketoacidosis.

These findings have major implications not only for patients with CKD and their families, but also health systems, given the enormous economic costs of managing kidney failure, especially the provision of dialysis and kidney transplant services.^18, 19^ A recent Kidney Health Australia commissioned report found the total annual cost of CKD was $9.9 billion in Australia in 2021.^20^ Early detection and best practice management of CKD, including implementation of SGLT2 inhibitors, was estimated to yield a net benefit of $10.2 billion in kidney failure and cardiovascular disease costs over the next two decades.^20^ Indeed, our results indicate that even modest increases in the uptake of SGLT2 inhibitors may have an important ramifications for the delivery of dialysis and transplant services in Australia.

Our findings build upon a growing body of literature that highlight the potential population health impacts of optimal implementation of this important drug class in people across the cardiorenal-metabolic spectrum. SGLT2 inhibitors have been shown to reduce the risk of heart failure hospitalisation and death from any cause in patients with heart failure, irrespective of ejection fraction or diabetes, and now form a central component of guideline-directed therapy for heart failure.^21^ Estimates from the United States indicate that almost 5 million patients with heart failure would be eligible for treatment with an SGLT2 inhibitor which, if optimally implemented, could prevent or postpone approximately 630,000 episodes of worsening heart failure or cardiovascular death over three years.^22^ If translated globally, between 7-8 million episodes of worsening heart failure or cardiovascular death could be averted.^23^ Similarly large numbers of patients with diabetes have been shown to be eligible for treatment with an SGLT2 inhibitor with considerable population-level benefits anticipated with optimal implementation.^24, 25^ Use of SGLT2 inhibitors has been shown to be cost-effective in economic evaluations across different settings and cardiometabolic diseases,^26, 27^ as reflected in the inclusion of SGLT2 inhibitors in the World Health Organisation List of Essential Medicines.^28^ Indeed compared to population wide approaches for preventing kidney failure due to diabetes such as lifestyle modification and sugar sweetened beverage taxes, modelling has demonstrated that widespread implementation of SGLT2 inhibitors would be the most effective strategy for reducing the incidence of kidney failure due to diabetes in Australia.^29^ Our results extend previous work to patients with CKD, regardless of diabetes, comparing eligibility for different agents within the medication class based on trial inclusion and exclusion criteria.

While the use of SGLT2 inhibitors is increasing modestly over time across some jurisdictions, most patients with CKD who would benefit from treatment are not receiving them in routine clinical practice, despite strong recommendations from major clinical practice guidelines advocating for their use. In fact, data from the United Kingdom and North America indicate that patients with CKD are less likely to receive an SGLT2 inhibitor than those with normal kidney function, despite the fact patients with CKD are at much higher risk of cardiovascular events and CKD progression and thus stand to gain more in absolute terms.^5, 30^ This ‘risk-treatment paradox’ is evident across the cardiorenal-metabolic spectrum, with people with established atherosclerotic cardiovascular disease also less likely to receive an SGLT2 inhibitor, despite greater absolute treatment benefits.^6, 31^ The reason for the underuse of SGLT2 inhibitors is complex and likely related to factors such as access and cost, perceived safety concerns, and therapeutic inertia to varying degrees. However, RAS blockade remains underused in people with CKD even two decades after landmark trials that established these medications as standard of care for patients with albuminuric CKD,^32^ suggesting that financial cost alone is not the only driver of underuse.

Taken together, the data highlight the urgency of efforts to maximise the implementation of SGLT2 inhibitors into routine clinical practice to reduce cardiorenal risk. How to do so is a priority area of research. The COORDINATE-Diabetes cluster randomized trial showed that a coordinated, multifaceted intervention of assessment, education and feedback improved the prescription of evidence-based therapies (SGLT2 inhibitors, GLP-1 receptor agonists and statins) in people with diabetes and atherosclerotic cardiovascular disease.^33^ In patients with heart failure, the STRONG-HF trial demonstrated that intensive post-discharge follow-up can optimise the prescription and dosing of guideline-directed heart failure therapy and reduce hospital readmission or all-cause death.^34^ COORDINATE-Diabetes and STRONG-HF highlight the need to evaluate implementation strategies to increase the uptake of proven therapies, including SGLT2 inhibitors, for patients with CKD to ensure that those who might benefit most from treatment are able to receive it.

This study has several strengths. Estimates of the number and proportion of patients with CKD eligible for treatment with an SGLT2 inhibitor were derived from a well characterized, nationally-representative cohort of patients in primary care. The study encompassed a contemporaneous cohort of patients with CKD in Australia reflecting the timeframe in which key SGLT2 inhibitor trials and major guideline updates were published. We estimated the impacts of improved uptake of SGLT2 inhibitors in a range of scenarios based on different CKD prevalence estimates and varying levels of treatment uptake.

Important limitations should be recognised when interpreting these findings. First, we applied distributions of sex- and age-stratified CKD prevalence in MedicineInsight to the broader Australian population based on national census data to estimate the number of patients with CKD in Australia. It is possible that there are differences in CKD prevalence in MedicineInsight relative to the broader Australian population. However, a previous study^35^ showed that CKD prevalence in MedicineInsight were similar when compared with other national estimates (Australia Health Survey), and we used the most conservative model of CKD prevalence possible (whereby prevalence of CKD was considerably less than that of other published estimates^20^). Second, as we did not have access to longer-term clinical outcomes (e.g., hospital admissions), we extrapolated event rates from SGLT2 inhibitor trials to the cohort, as has been done in analogous work in diabetes and in heart failure.^22–24^ However, these event rates may not be applicable to routine practice due to differences in baseline risk. Indeed, we observed important differences in patient characteristics (including eGFR and UACR) and use of RAS blockade between these groups. Treatment adherence in routine practice is typically lower than clinical trials and may wane over time. These factors may reduce the estimated population-level impacts of improved SGLT2 inhibitor uptake. Finally, we were unable to apply some exclusion criteria (e.g., polycystic kidney disease) as we did not have data on CKD aetiology. However, the impact of this is likely minimal given these conditions occur uncommonly at a population level.

In conclusion, we estimate that a substantial number of patients with CKD in Australia are eligible for treatment with SGLT2 inhibitors. Optimal implementation of SGLT2 inhibitors for patients with CKD is projected to translate to substantial population-level benefits including potentially preventing large numbers of patients experiencing CKD progression or dying due to cardiovascular disease or kidney failure. Identifying strategies to increase the uptake of SGLT2 inhibitors in patients with CKD is critical to realising the population-level benefits of this drug class.

## Author contributions

BLN, MJ, JW, SK, SVB, MG and PER contributed to the concept and rationale for the study and interpretation of the results. KN and MJ were responsible for acquisition of the data extract from MedicineInsight. BLN, MJ, JW and PER developed the study protocol and oversaw the implementation of the study analytical plan. BLN drafted the initial manuscript. All authors contributed to the design of the study, interpretation of the data and critical revision of the manuscript.

## Data Availability

The current study is based on data from MedicineInsight, a national general practice data source developed by NPS MedicineWise and managed by the Australian Commission on Safety and Quality in Health Care. The 2021 Australian population census data is publicly available via the Australian Bureau of Statistics website (www.abs.gov.au). All relevant data are within the manuscript and its supplementary appendix.

## Acknowledgements

This study is based on data from MedicineInsight (project 2020-004), a national general practice data program developed by NPS MedicineWise and managed by the Australian Commission on Safety and Quality in Health Care. MedicineInsight extracts and collates longitudinal, de-identified, patient health data from the clinical information systems of consenting general practices across Australia. This study was supported by an unrestricted research grant from Boehringer Ingelheim.

## Supplementary appendix

**Figure S1.**
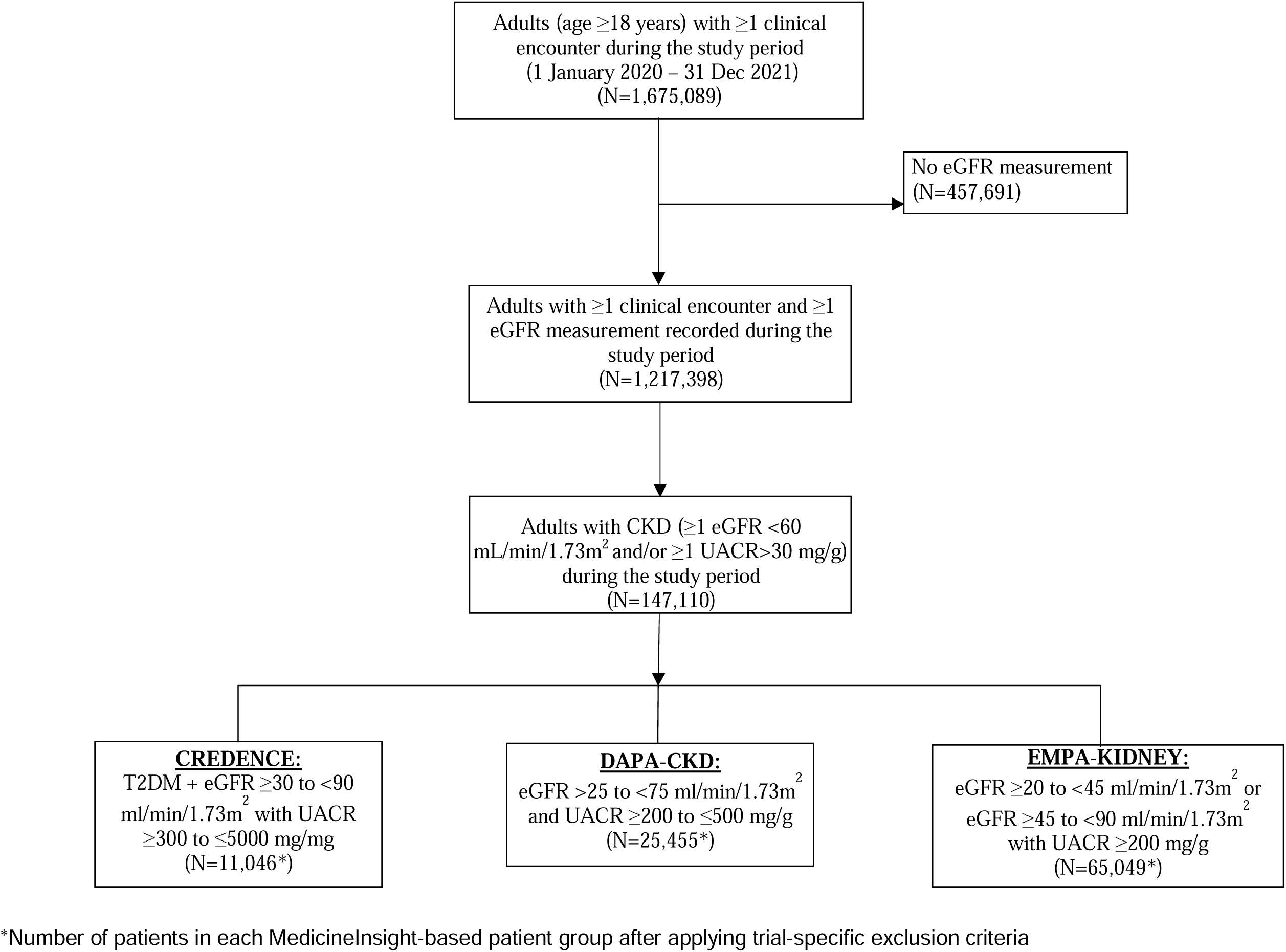
Identification of the study cohort in MedicineInsight

**Figure S2.**
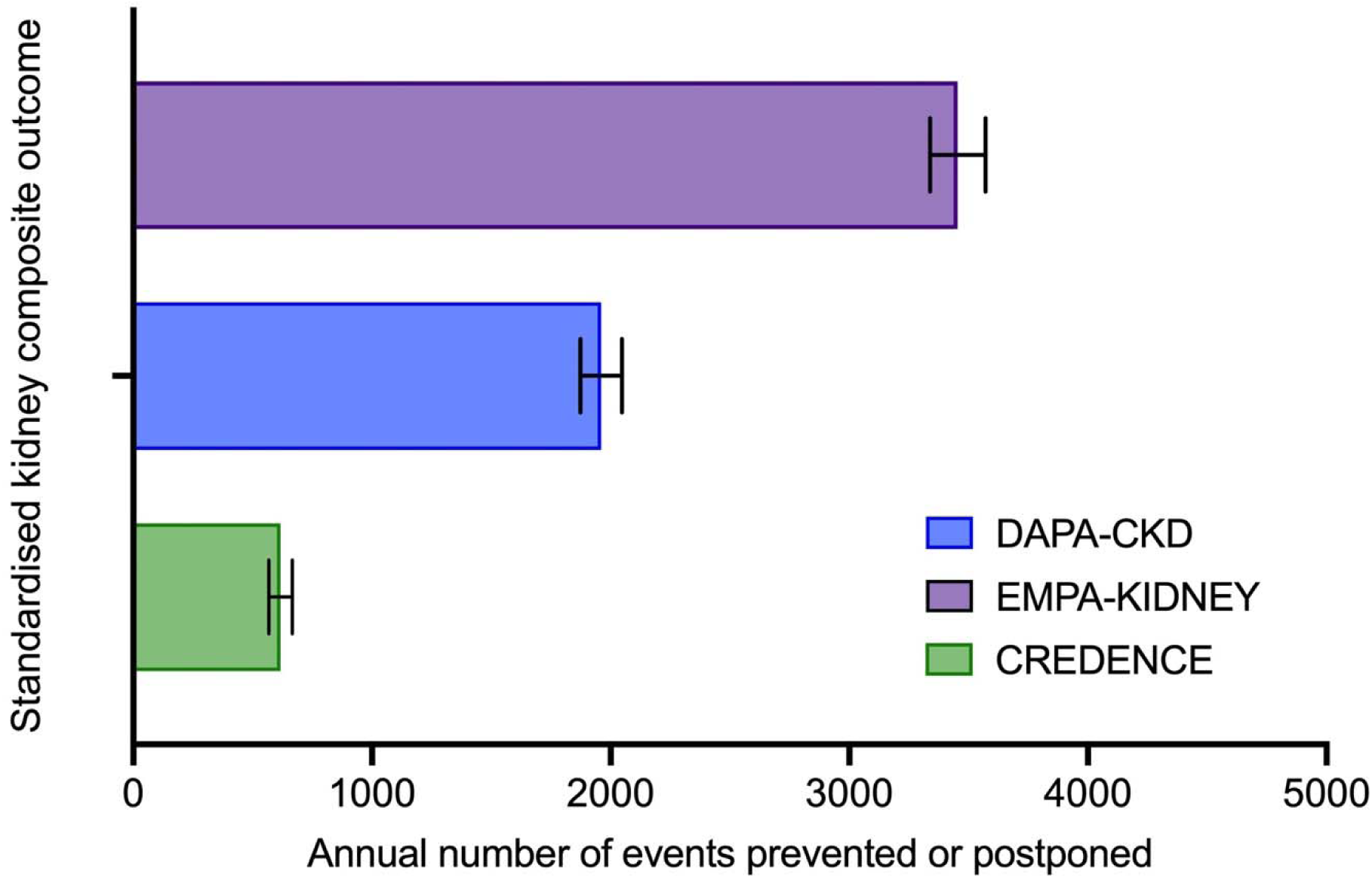
Estimated annual number of events potentially prevented with optimal implementation (75% uptake) of SGLT2 inhibitors in patients with CKD in Australia using a standardised kidney composite outcome of sustained 40% decline in eGFR, kidney failure, or death due to kidney failure.

**Table S1.** Trial-specific inclusion/exclusion criteria

|  | **EMPA-KIDNEY** |
| --- | --- |
| Inclusion criteria | 1. Aged $\geq 18$ years<br>2. Evidence of progressive CKD at risk of kidney disease progression is defined on the basis of local laboratory results recorded at least 3 months before and at the time of the Screening visit, and requires that:<br>(a) CKD-EPI eGFR $\geq 20$ and $< 45$ mL/min/1.73m<sup>2</sup> or<br>(b) CKD-EPI eGFR $\geq 45$ and $< 90$ mL/min/1.73m<sup>2</sup> with uACR $\geq 200$ mg/g (or protein:creatinine ratio $\geq 300$ mg/g)<br>3. A local investigator judges that the participant neither requires empagliflozin (or any other SGLT-2 or SGLT-1/2 inhibitor), nor that such treatment is definitely inappropriate<br>4. Treated with appropriate doses of RAS-inhibition, unless such treatment is either not tolerated or not indicated. |
| Exclusion criteria | 1. Currently receiving SGLT-2 or SGLT-1/2 inhibitor<br>2. Diabetes mellitus type 2 and prior atherosclerotic cardiovascular disease with an eGFR $> 60$ mL/min/1.73m<sup>2</sup> at screening<br>3. Receiving combined ACEi and ARB treatment<br>4. Maintenance dialysis, functioning kidney transplant, or scheduled living donor transplant*; (v) Polycystic kidney disease<br>5. Previous or scheduled bariatric surgery<br>6. Ketoacidosis in the past 5 years<br>7. Symptomatic hypotension, or systolic blood pressure $< 90$ or $> 180$ mmHg at Screening<br>8. ALT or AST $> 3 \times$ ULN at screening<br>9. Hypersensitivity to empagliflozin or another SGLT2 inhibitor<br>10. Any intravenous immunosuppression therapy in last 3 months; or anyone currently on $> 45$ mg prednisolone (or equivalent)<br>11. Use of an investigational medicinal product in the 30 days prior to screening visit<br>12. Known to be poorly compliant with clinic visits or prescribed medication<br>13. Medical history that might limit the individual's ability to take trial treatments for the duration of the study (e.g. severe respiratory disease; history of cancer or evidence of spread within last 4 years, other than non-melanoma skin cancer; or recent history of alcohol or substance misuse)<br>14. Current pregnancy, lactation or women of childbearing potential, unless using highly- effective contraception<br>15. Type 1 diabetes mellitus (introduced in Protocol version 2.0)<br>In addition, individuals will be excluded at the Randomization visit if the participant:<br>1. Does not adhere to run-in treatment<br>2. Is no longer willing to be randomized and followed for at least 3 years<br>3. Is considered by a local investigator not to be suitable for randomization or<br>4. Experiences ketoacidosis, heart attack, stroke, or hospitalization for heart failure, or hospitalization for urinary tract infection or acute kidney injury during run-in |

|  | **DAPA-CKD** |
| --- | --- |
| Inclusion criteria | 1. Provision of signed informed consent prior to any study specific procedures<br>2. Female or male aged $\geq 18$ years at the time of consent<br>3. eGFR $\geq 25$ and $\leq 75$ mL/min/1.73m<sup>2</sup> (CKD-EPI Formula) at visit 1<br>4. UACR $\geq 200$ and $\leq 5000$ mg/g at visit 1<br>5. Stable, and for the patient maximum tolerated labelled daily dose, treatment |
|  | with angiotensin converting enzyme (ACE) inhibitor or angiotensin receptor blocker (ARB) for at least 4 weeks before visit 1, if not medically contraindicated |
| Exclusion criteria | 1. Autosomal dominant or autosomal recessive polycystic kidney disease, lupus nephritis or anti-neutrophil cytoplasmic antibody (ANCA)-associated vasculitis<br>2. Receiving cytotoxic therapy, immunosuppressive therapy or other immunotherapy for primary or secondary renal disease within 6 months prior to enrolment<br>3. History of organ transplantation<br>4. Receiving therapy with a sodium glucose co-transporter 2 (SGLT2) inhibitor within 8 weeks prior to enrolment or previous intolerance of an SGLT2 inhibitor<br>5. Type 1 diabetes<br>6. New York Heart Association (NYHA) class IV Congestive Heart Failure at the time of enrolment<br>7. MI, unstable angina, stroke or transient ischemic attack (TIA) within 12 weeks prior to enrolment<br>8. Coronary revascularisation (percutaneous coronary intervention [PCI] or coronary artery bypass grafting [CABG]) or valvular repair/replacement within 12 weeks prior to enrolment or is planned to undergo any of these procedures after randomization<br>9. Any condition outside the renal and cardiovascular disease area, such as but not limited to malignancy, with a life expectancy of less than 2 years based on investigator's clinical judgement<br>10. Active malignancy requiring treatment at the time of visit 1 (with the exception of successfully treated basal cell or treated squamous cell carcinoma).<br>11. Hepatic impairment (aspartate transaminase [AST] or alanine transaminase [ALT] >3x the upper limit of normal [ULN]; or total bilirubin >2x ULN at time of enrolment)<br>12. Known blood-borne diseases<br>13. Women of child-bearing potential (ie, those who are not chemically or surgically sterilized or who are not post-menopausal) who are not willing to use a medically accepted method of contraception that is considered reliable in the judgment of the investigator OR women who have a positive pregnancy test at enrolment or randomisation OR women who are breast-feeding<br>14. Participation in another clinical study with an investigational product (IP) during the last month prior to Enrolment<br>15. Inability of the patient, in the opinion of the investigator, to understand and/or comply with IP, procedures and/or follow-up OR any conditions that, in the opinion of the investigator, may render the patient unable to complete the study. |
|  | **CREDENCE** |
| Inclusion criteria | 1. Man or woman ≥30 years-old with a clinical diagnosis of type 2 diabetes mellitus (T2DM)<br>2. Glycated hemoglobin (HbA1c) ≥6.5% to ≤12.0%<br>3. Estimated glomerular filtration rate (eGFR) ≥30 to <90 mL/min/1.73 m<sup>2</sup> (as determined using the Chronic Kidney Disease Epidemiology Collaboration [CKD-EPI] equation)<br>4. Urinary albumin:creatinine ratio (UACR) >300 mg/g to ≤5000 mg/g (>33.9 mg/mmol to ≤565.6 mg/mmol)<br>5. All subjects must be on a stable maximum tolerated labelled daily dose of ACEi or ARB for at least 4 weeks prior to randomization |
|  | 6. Women must be:<br>i. Postmenopausal, defined as:<br>a. >45 years of age with amenorrhea for at least 18 months, or<br>b. >45 years of age with amenorrhea for at least 6 months and <18 months and a serum follicle stimulating hormone (FSH) level >40 IU/L, or<br>c. Surgically sterile (have had a hysterectomy or bilateral oophorectomy, tubal occlusion or otherwise be incapable of pregnancy, or<br>d. Heterosexually active and practicing a highly effective method of birth control or<br>e. not heterosexually active<br>ii. Women of childbearing potential (i.e., those subjects who do not meet the postmenopausal definition above), regardless of age, must have a negative urine pregnancy test at baseline and at screening if required by local regulations<br>7. Willing and able to adhere to the prohibitions and restrictions specified in this protocol<br>8. Subjects must have signed an informed consent document indicating that they understand the purpose of and procedures required for the study and are willing to participate in the study<br>9. Inclusion Criterion for Randomization:<br>i. Subjects must have ≥80% compliance (by pill count) with single-blind placebo |
| Exclusion criteria | 1. History of diabetic ketoacidosis or type 1 diabetes mellitus<br>2. History of hereditary glucose-galactose malabsorption or primary renal glucosuria<br>3. Known medical history or clinical evidence suggesting nondiabetic renal disease<br>4. Renal disease that required treatment with immunosuppressive therapy or a history of chronic dialysis or renal transplant<br>5. Uncontrolled hypertension (systolic blood pressure [BP] ≥180 and/or diastolic BP ≥100 mmHg) by Week -2<br>6. Blood potassium level >5.5 mmol/L during screening<br>7. Myocardial infarction, unstable angina, revascularization procedure (e.g., stent or bypass graft surgery), or cerebrovascular accident within 12 weeks before randomization, or a revascularization procedure is planned during the trial<br>8. Current or history of heart failure of New York Heart Association (NYHA) class IV cardiac disease<br>9. Electrocardiogram (ECG) findings within 12 weeks before randomization that would require urgent diagnostic evaluation or intervention (e.g., new clinically important arrhythmia or conduction disturbance)<br>10. Known significant liver disease (e.g., acute hepatitis, chronic active hepatitis, cirrhosis)<br>11. Alanine aminotransferase (ALT) levels >2.0 times the upper limit of normal (ULN) or total bilirubin >1.5 times the ULN, unless in the opinion of the investigator and as agreed upon by the sponsor's medical officer, the findings are consistent with Gilbert's disease<br>12. History of malignancy within 5 years before screening (exceptions: squamous and basal cell carcinomas of the skin and carcinoma of the cervix in situ, or a malignancy that in the opinion of the investigator, with concurrence with the sponsor's medical monitor, is considered cured with minimal risk of recurrence)<br>13. History of human immunodeficiency virus (HIV) antibody positive<br>14. Major surgery within 12 weeks before randomization, or has not fully recovered from surgery<br>15. Any condition that in the opinion of the investigator or sponsor's medical monitor would make participation not in the best interest of the subject, or could prevent, limit, or confound the protocol- specified assessments<br>16. History of atraumatic amputation within past 12 months of screening, or an active skin ulcer, osteomyelitis, gangrene, or critical ischemia of the lower extremity |
|  | within 6 months of screening (added May 5, 2016)<br>17. Combination use of an ACEi and ARB<br>18. Use of an MRA or a direct renin inhibitor (DRI)<br>19. Current use of a sodium glucose co-transporter 2 (SGLT2) inhibitor (within 12 weeks prior to randomization)<br>20. Current participation in another canagliflozin study or previously exposed to canagliflozin in a prior canagliflozin study<br>21. Known allergies, hypersensitivity, or intolerance to canagliflozin or its excipients<br>22. Received an active investigational drug (including vaccines) other than a placebo agent, or used an investigational medical device within 12 weeks before Day 1/baseline<br>23. Pregnant or breast-feeding or planning to become pregnant or breast-feed during the study |

**Table S2.** Prevalence of CKD in 2020 by sex and age group in MedicineInsight*

| Sex | Age group (years) | Overall number of patients in the study cohort | High-estimate model |  | Moderate model |  | Conservative model |  |
| --- | --- | --- | --- | --- | --- | --- | --- | --- |
|  |  |  | Number | % | Number | % | Number | % |
| Females |  |  |  |  |  |  |  |  |
|  | 18-29 | 141,031 | 472 | 0.3 | 407 | 0.3 | 62 | 0.1 |
|  | 30-39 | 139,717 | 993 | 0.7 | 809 | 0.6 | 162 | 0.1 |
|  | 40-49 | 132,687 | 1,934 | 1.5 | 1,459 | 1.1 | 387 | 0.3 |
|  | 50-59 | 138,557 | 4,317 | 3.1 | 2,750 | 2.0 | 994 | 0.7 |
|  | 60-69 | 129,216 | 9,516 | 7.4 | 5,169 | 4.0 | 2,710 | 2.1 |
|  | 70-79 | 101,792 | 20,110 | 19.8 | 10,159 | 10.0 | 7,425 | 7.3 |
|  | ≥80 | 67,875 | 27,662 | 40.8 | 13,398 | 19.7 | 11,437 | 16.9 |
| Males |  |  |  |  |  |  |  |  |
|  | 18-29 | 73,859 | 327 | 0.4 | 266 | 0.4 | 61 | 0.1 |
|  | 30-39 | 82,103 | 864 | 1.1 | 718 | 0.9 | 173 | 0.2 |
|  | 40-49 | 97,506 | 1,982 | 2.0 | 1,537 | 1.6 | 432 | 0.4 |
|  | 50-59 | 112,455 | 4,662 | 4.2 | 3,315 | 3.0 | 1,169 | 1.0 |
|  | 60-69 | 113,288 | 10,651 | 9.4 | 6,685 | 5.9 | 3,407 | 3.0 |
|  | 70-79 | 90,965 | 20,046 | 22.0 | 11,462 | 12.6 | 7,884 | 8.7 |
|  | ≥80 | 48,782 | 20,248 | 41.5 | 10,732 | 22.0 | 8,899 | 18.2 |

\*Data from study which assessed the prevalence of CKD in MedicineInsight<sup>14</sup>.

**Table S3.**
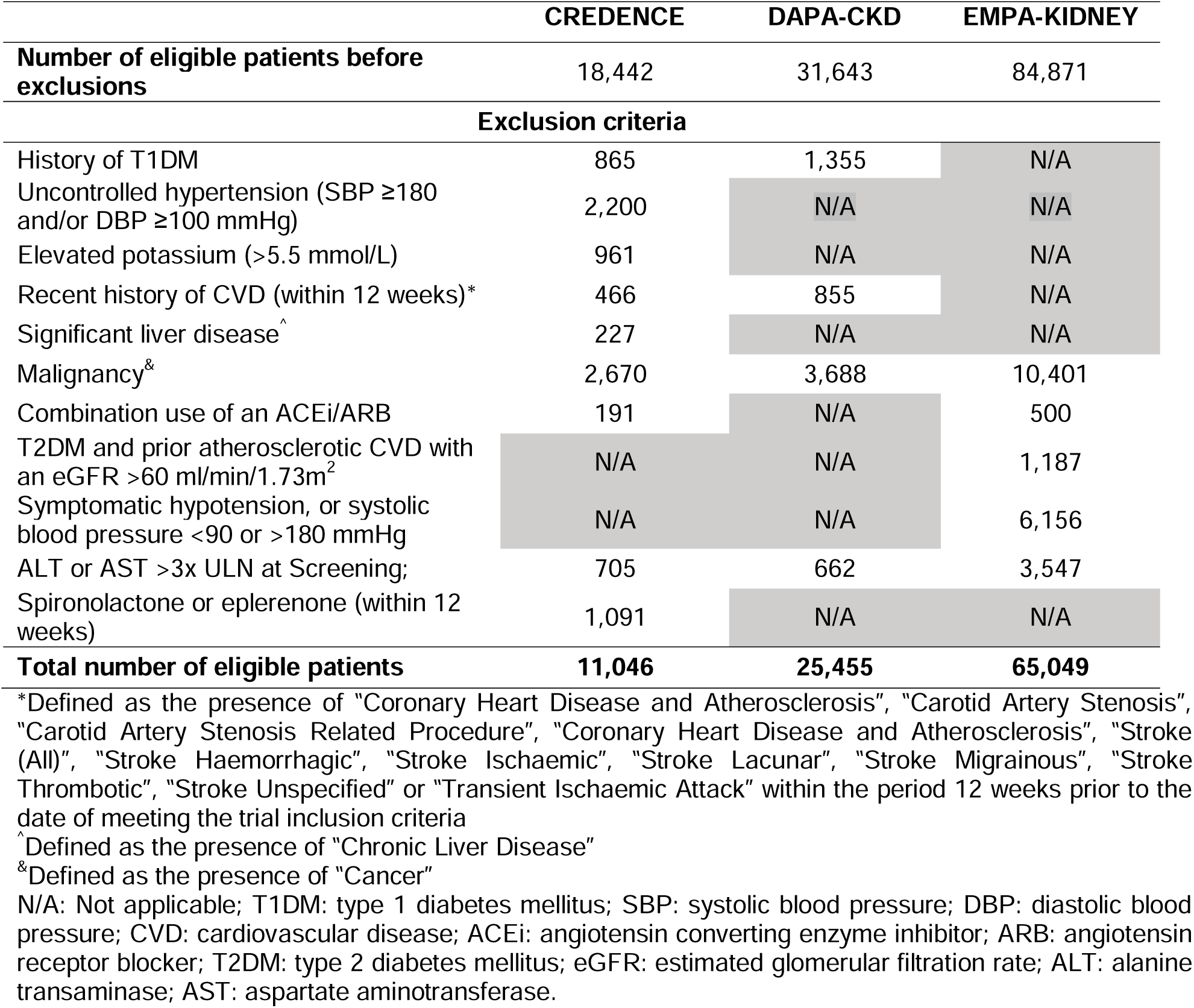
Number of patients with CKD in MedicineInsight excluded from key SGLT2 inhibitor kidney outcome trials

**Table S4.**
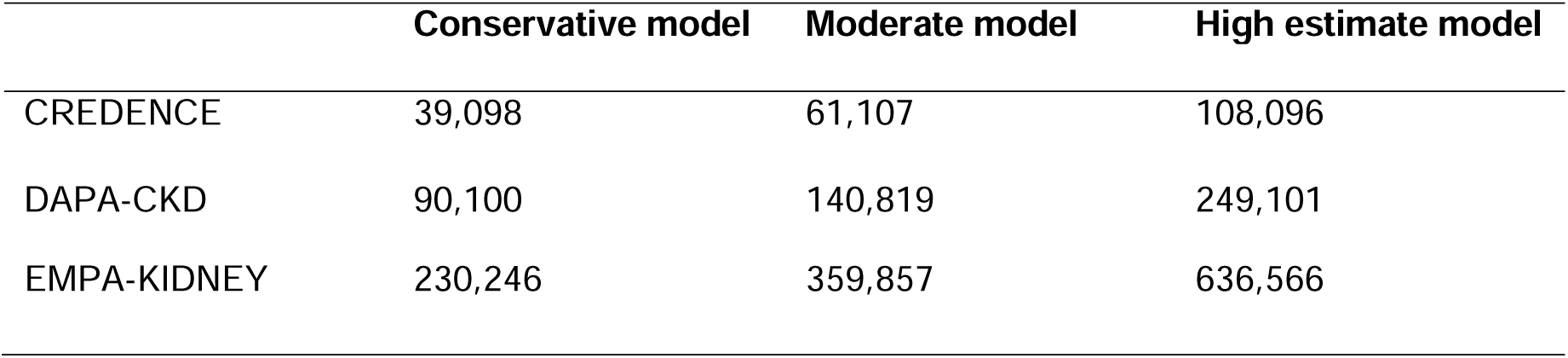
Estimated number of patients with CKD in Australia who would have been eligible for treatment with an SGLT2 inhibitor based on different estimates of CKD prevalence.

**Table S5.**
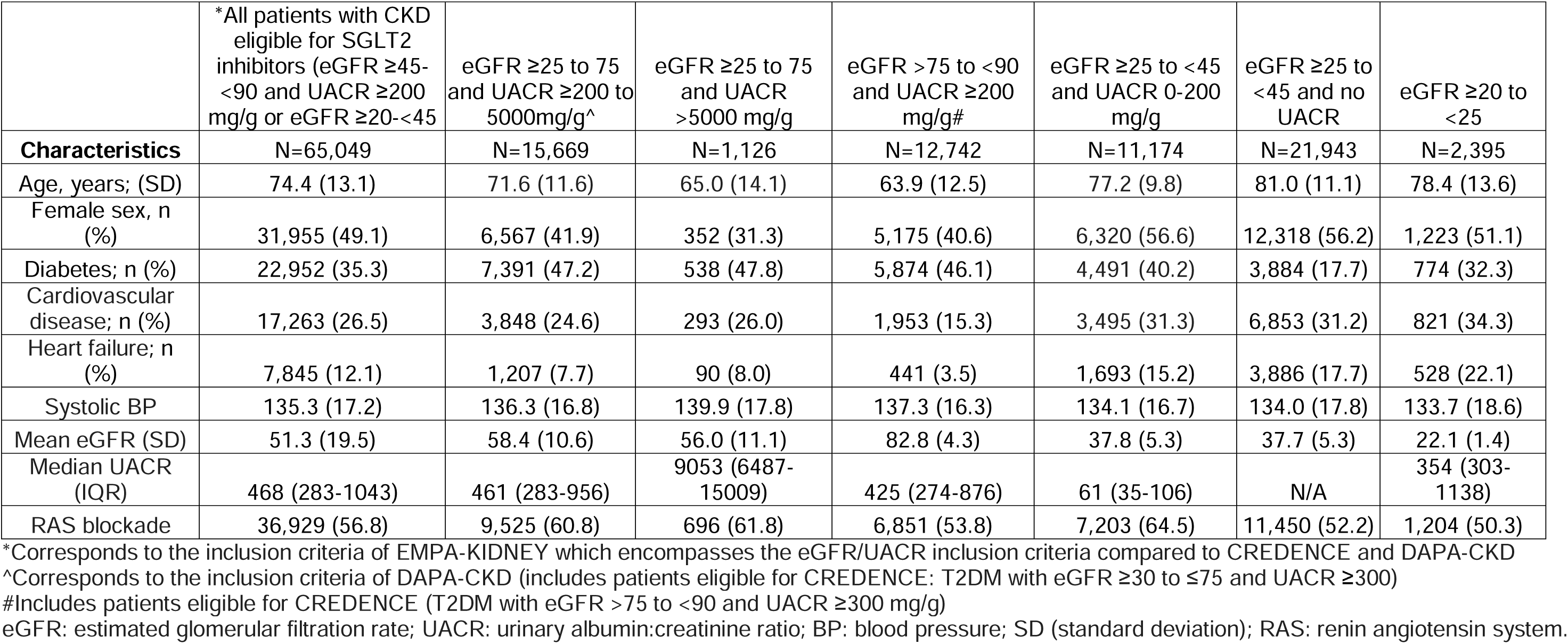
Characteristics of eligible populations in CREDENCE, DAPA-CKD and EMPA-KIDNEY.

**Table S6.**
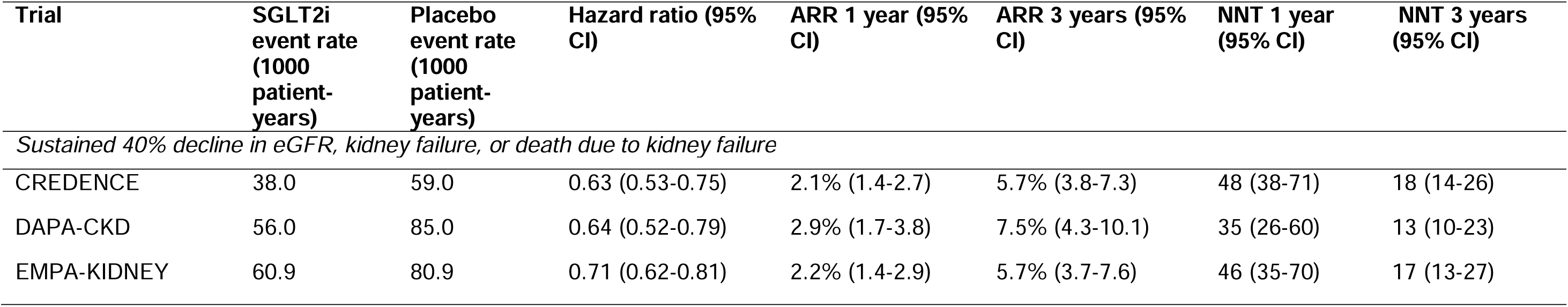
Event rates, treatment effects and absolute risk reductions at 1 and 3 years for the standardised kidney composite outcome of sustained 40% decline in eGFR, kidney failure or death due to kidney failure.

**Table S7.**
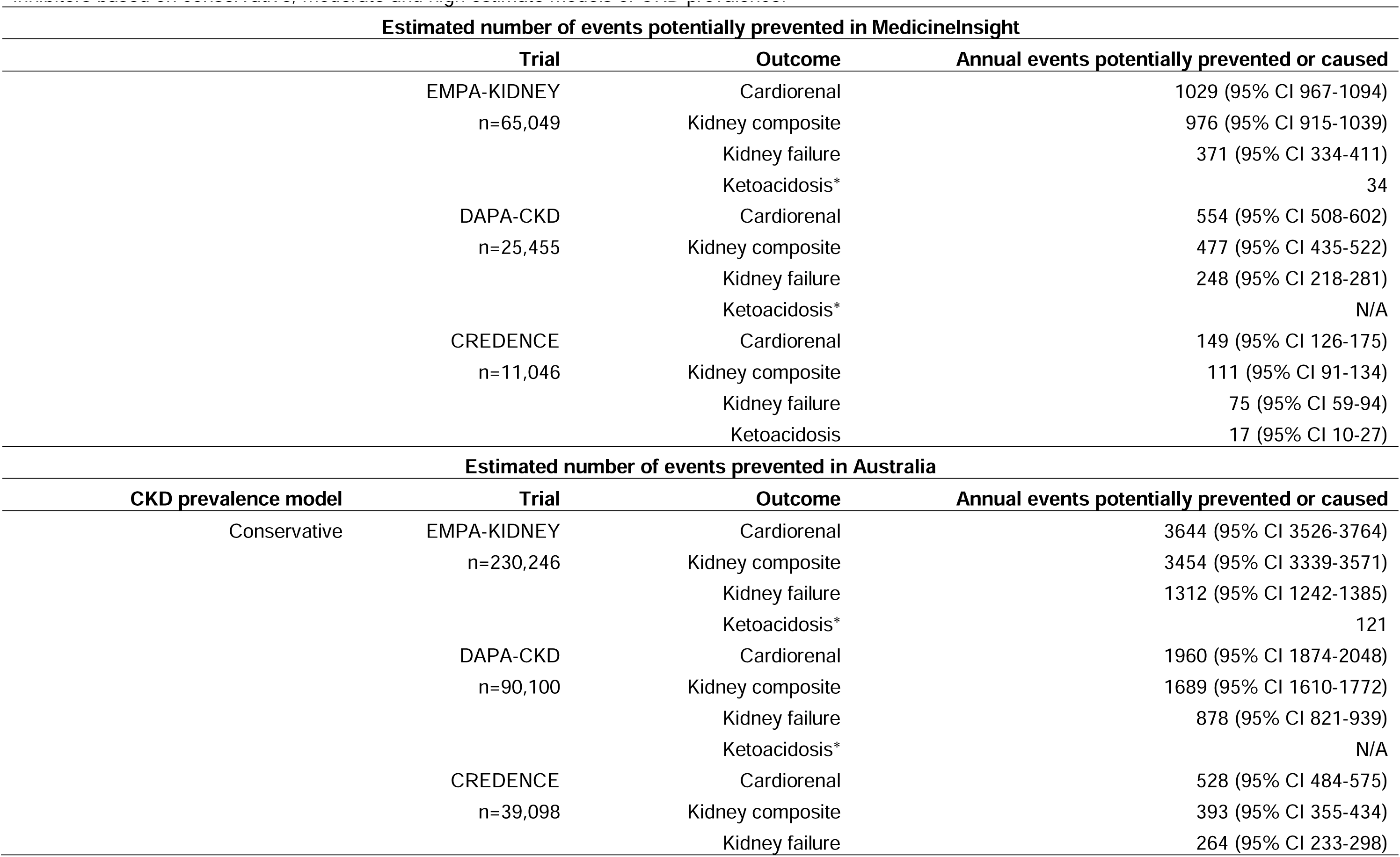
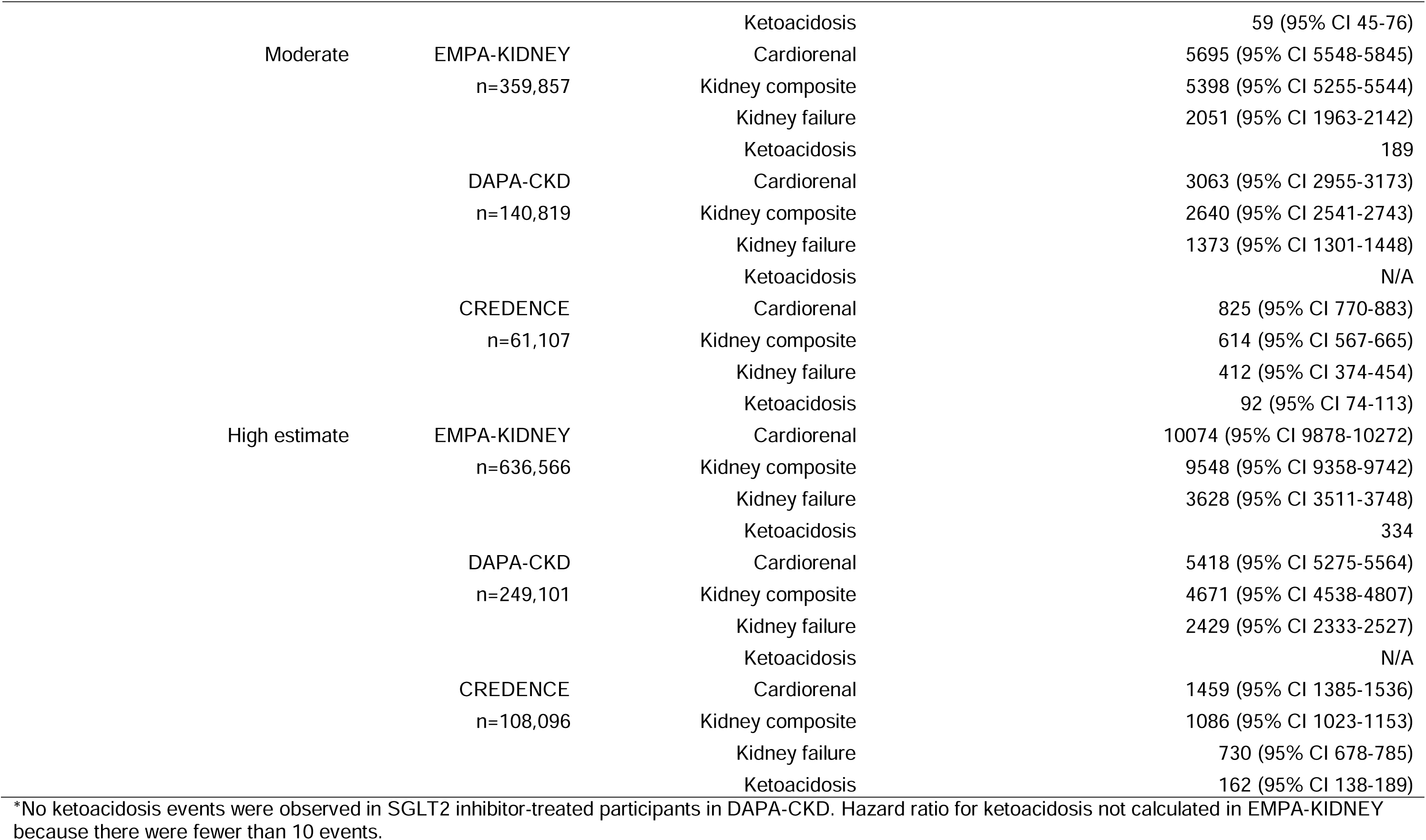
Estimated number of events potentially prevented or caused in MedicineInsight and in Australia with optimal implementation (75% uptake) of SGLT2 inhibitors based on conservative, moderate and high estimate models of CKD prevalence.

**Table S8.** Number of events, event rates, relative treatment effects for the primary cardiorenal outcome across diabetes, eGFR and UACR subgroups.

|  | Events (n/N) |  | Event rate (1000 patient-years)* |  |  |
| --- | --- | --- | --- | --- | --- |
| Trial/Subgroup | SGLT2i | Placebo | SGLT2i | Placebo | HR (95% CI) |
| EMPA-KIDNEY |  |  |  |  |  |
| Diabetes | 218/1525 | 306/1515 | 71.5 | 101.0 | 0.64(0.54-0.77) |
| No diabetes | 214/1779 | 252/1790 | 60.1 | 70.4 | 0.82(0.68-0.99) |
| eGFR |  |  |  |  |  |
| <45 mL/min/1.73m<sup>2</sup> | 387/2598 | 492/2612 | 74.5 | 94.2 | 0.64(0.44-0.93) |
| ≥45 mL/min/1.73m<sup>2</sup> | 45/706 | 66/693 | 31.9 | 47.6 | 0.75(0.66-0.85) |
| UACR |  |  |  |  |  |
| ≤300 mg/g | 109/1592 | 120/1600 | 31.9 | 47.6 | 0.95(0.73-1.23) |
| >300 mg/g | 323/1712 | 438/1705 | 94.3 | 128.4 | 0.67(0.58-0.78) |
| DAPA-CKD |  |  |  |  |  |
| Diabetes | 152/1455 | 229/1451 | 52.0 | 80.0 | 0.64(0.52-0.79) |
| No diabetes | 45/697 | 83/701 | 34.0 | 63.0 | 0.50(0.35-0.72) |
| eGFR |  |  |  |  |  |
| <45 mL/min/1.73m<sup>2</sup> | 152/1455 | 229/1451 | 49.8 | 72.3 | 0.63(0.51-0.78) |
| ≥45 mL/min/1.73m<sup>2</sup> | 45/697 | 83/701 | 21.3 | 38.3 | 0.49(0.34-0.69) |
| UACR |  |  |  |  |  |
| ≤300 mg/g | 9/240 | 9/205 | 18.0 | 21.0 | 0.86(0.34-2.16) |
| >300 mg/g | 188/1912 | 303/1947 | 50.0 | 81.0 | 0.61(0.51-0.73) |
| **CREDENCE** |  |  |  |  |  |
| <45 mL/min/1.73m<sup>2</sup> | 122/679 | 166/687 | 71.9 | 99.9 | 0.69(0.55-0.86) |
| ≥45 mL/min/1.73m<sup>2</sup> | 123/1523 | 174/1512 | 31.0 | 44.7 | 0.70(0.59-0.82) |
| UACR** |  |  |  |  |  |
| ≤300 mg/g | - | - | - | - | - |
| >300 mg/g | 245/2202 | 340/2199 | 43.2 | 61.2 | 0.70(0.59-0.82) |

\*Event rates estimated based on number of events and median follow-up duration for EMPA-KIDNEY and for DAPA-CKD eGFR subgroups. SGLT2: sodium glucose cotransporter 2 inhibitor; eGFR: estimated glomerular filtration rate; UACR: urinary albumin:creatinine ratio.

**Table S9.**
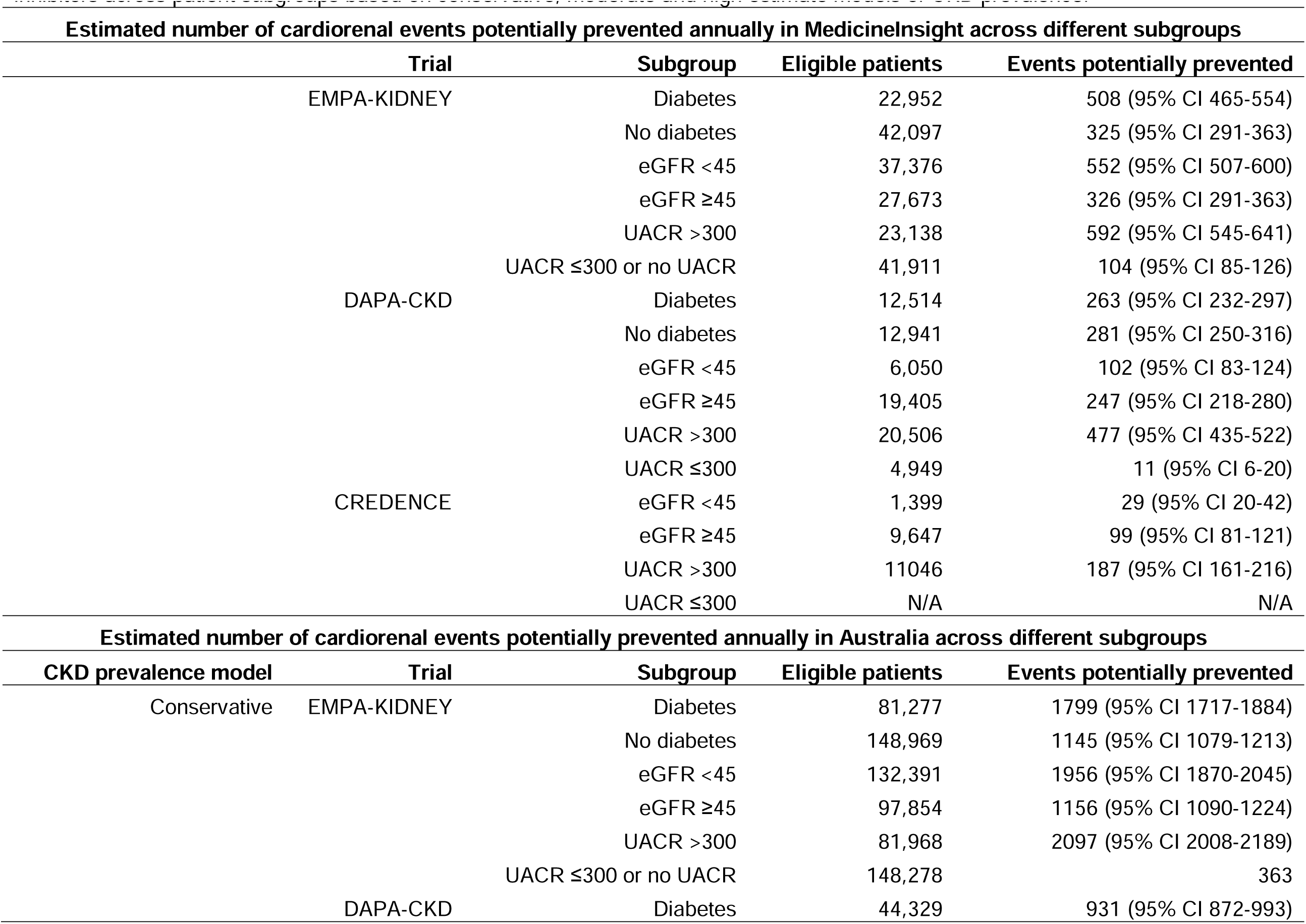
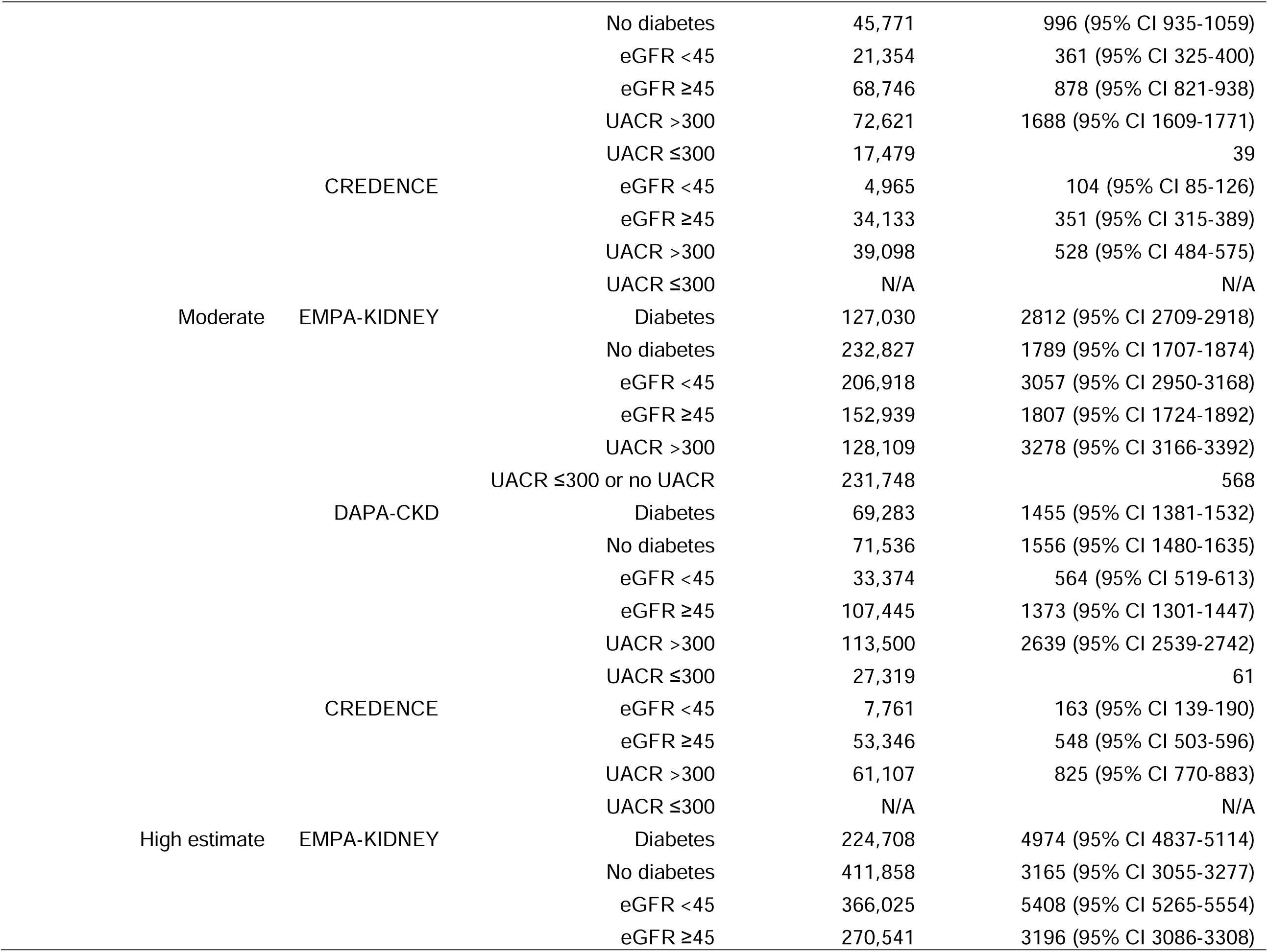
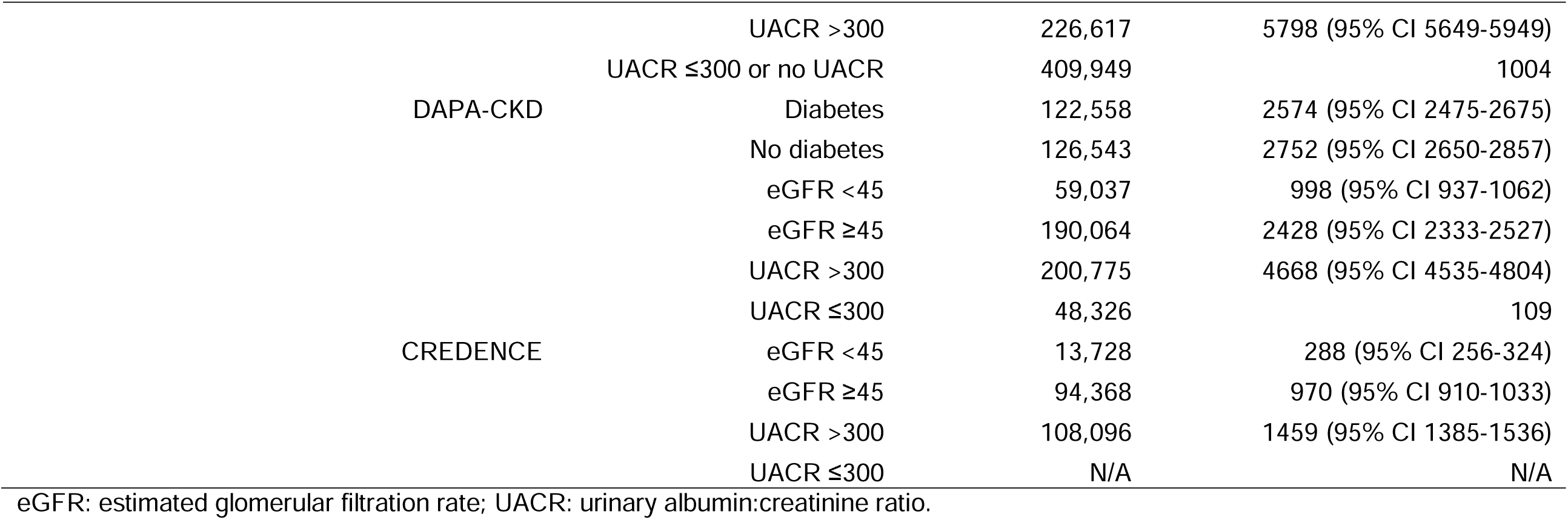
Estimated number of events potentially prevented or caused in MedicineInsight and in Australia with optimal implementation (75% uptake) of SGLT2 inhibitors across patient subgroups based on conservative, moderate and high estimate models of CKD prevalence.

